# Impact of clinical severity on plasma p-tau performance in predementia Alzheimer’s disease

**DOI:** 10.1101/2024.08.06.24311532

**Authors:** Fernando Gonzalez-Ortiz, Bjørn-Eivind Kirsebom, Yara Yakoub, Julia K. Gundersen, Lene Pålhaugen, Knut Waterloo, Per Selnes, Jonas Alexander Jarholm, Berglind Gísladóttir, Arvid Rongve, Ragnhild Eide Skogseth, Geir Bråthen, Dag Aarsland, Michael Turton, Peter Harrison, Henrik Zetterberg, Sylvia Villeneuve, PREVENT AD research group, Tormod Fladby, Kaj Blennow

## Abstract

**Background and objectives:** Detecting Alzheimer’s disease (AD) biological and clinical changes is crucial for early diagnostic and therapeutic interventions. Here we investigate the relationship between clinical severity and levels of phosphorylated tau, focusing on plasma biomarkers, in preclinical and prodromal AD.

**Methods:** In this study (n=621), we examined two independent cohorts consisting of preclinical and prodromal AD. Cohort-1 included 431 participants classified as either cognitively normal (CN) or mild cognitive impaired (MCI) with normal or abnormal cerebrospinal fluid (CSF) Aβ42/40 ratio (A) and p-tau181 (T) [CN A-/T-, n=169; A+/T-, CN=26; MCI=24; A+/T+, CN=40; MCI=105; CN=34; MCI=33]. A total of n=418 of the participants had longitudinal assessments of verbal memory up to 9.67 years from baseline. Cohort-2 included 190 participants in whom amyloid status was determined using Aβ positron emission tomography (PET) [Aβ- CN= 118; Aβ+ CN= 49; Aβ+ MCI= 21].

**Results:** In cohort-1, plasma p-tau217 showed a moderate correlation with its corresponding CSF biomarker (rho=0.65, p<.001) and high accuracy identifying Aβ+ participants (AUC: 0.85). Diagnostic accuracy of plasma p-tau217 was significantly greater for MCI Aβ+ (AUC: 0.89) versus CN Aβ+ (AUC: 0.79, p<.05) and for A+/T+ (AUC: 0.88) versus A+/T- (AUC: 0.78, p<.05). P-tau181 and p-tau231 showed significantly weaker CSF-plasma correlations (rho= 0.47, and rho=0.32, p<.001, respectively) and levels were not as tightly associated with cognitive status in the Aβ+ group. While all the CSF p-tau markers were associated to future cognitive deterioration, p-tau217 was the only plasma biomarker associated with future memory decline (β=0.05, p<0.05). In cohort 1 and 2, plasma p-tau217 showed significantly higher concentrations in MCI Aβ+ as compared to CN Aβ+. Furthermore, plasma p-tau217 demonstrates similar biomarker elevations when compared to CN Aβ- controls in both cohorts.

**Discussion:** Our findings indicate that, unlike p-tau181 and p-tau231, plasma p-tau217 consistently aligns with cognitive status in Aβ+ individuals and more closely reflects CSF biomarker abnormalities, potentially reducing discrepancies between clinical and biochemical findings. The associations of plasma p-tau217 with baseline and future cognitive decline make it a valuable complement to clinical evaluations in preclinical and prodromal AD, especially when CSF analysis or PET are not feasible.

## Introduction

Alzheimer’s disease (AD) represents the most common cause of dementia in the elderly^1^. However, despite its high prevalence, a clinical diagnosis of AD remains challenging due to its insidious symptomatology, particularly in early stages of the disease^2–4^. While definite diagnosis is based on the neuropathological evidence of the main AD hallmarks, ß-amyloid plaques and tau tangles in the brain^1,5^, the use of neuroimaging (positron emission tomography [PET]) and cerebrospinal fluid (CSF) biomarkers that corroborate the presence of these hallmarks has proven to be valuable to support clinical diagnosis^1,6,7^. These pathological changes are often detected in the brain well before any noticeable cognitive change, reflecting the preclinical or prodromal stages^2,8^, which may explain why biologically defined AD is more prevalent than clinically defined AD^9^

Classification or staging of AD pathology can be assessed through biological and cognitive measurements. Biological classification relies on objective markers identified through imaging or CSF analysis (e.g., amyloid PET and CSF amyloid42/40 ratio). These biomarkers provide valuable insights into the disease at the molecular level and categorize patients according to the presence of amyloid and tau pathology^10^. On the other hand, clinical classification of AD involves the assessment of cognitive and functional decline^11^.

In recent years, sensitive techniques for assessing biomarkers in plasma have emerged as accessible methods for detecting and potentially predicting early AD changes^6,12^. Plasma p-tau markers such as p-tau181, p-tau217 and p-tau231 have shown promising performance to identify patients with early amyloid pathology^7,8,12^. Among these markers, p-tau217 has shown a superior accuracy in early stages of the disease continuum, serving as a possible first-in-line diagnostic test^13,14^.

While plasma biomarkers hold great promise for early AD diagnosis and prognosis^15^, they do not consistently align with the stage of cognitive decline^16^. Some individuals with substantial biomarker evidence of AD pathology may exhibit mild clinical symptoms, whereas others with fewer biomarker abnormalities may experience more severe cognitive deficits^3,16^. Moreover, the temporal evolution of AD biomarkers is not linearly correlated with cognitive decline^17,18^. The rate of change in biomarker levels may vary greatly between individuals, and clinical symptoms may worsen rapidly or slowly, making it challenging to predict disease progression accurately^17^. The reasons for this are not completely understood but may relate to the temporal disconnect between biomarker evidence of pathology build-up and neuronal network breakdown/loss of brain resilience^19^.

In this study, we investigate changes in p-tau biomarkers, with a focus on plasma, in relation to clinical severity in predementia AD. We also explore their prognostic capabilities by examining associations with cognitive status and future cognitive deterioration in early AD. Additionally, we compare the positive predictive values (PPVs) and negative predictive values (NPVs) of each plasma p-tau markers to determine their precision and appropriateness of use at different clinical stages of predementia AD.

## Methods

### Standard Protocol Approvals, Registrations, and Patient Consents

The present study was performed with ethical pertinent ethical approvals. Cohort 1 (DDI) has been approved by the Regional Committees for Medical and Health Research Ethics in Norway (REK: 2013/150)^20^. All participants gave a written informed consent before participating in the study. A detailed description of inclusion and exclusion criteria are outlined in Fladby et al., (2017). Cohort-2 (PREVENT-AD) included Written informed consent was obtained from all participants and all research procedures were approved by the Institutional Review Board at McGill University.

### Cohort-1: Dementia Disease Initiation (DDI)

The DDI study is a Norwegian multicentre cohort recruiting participants across all university hospitals in Norway. See Fladby et al. (2017)^20^ for details. The DDI cohort includes predementia cases with either Subjective Cognitive Decline (SCD) or MCI staged according to published criteria ^21,22^ and participants recruited as controls primarily from spouses of recruited patients, and secondarily from advertisements in local news media and also orthopaedic patients who had lumbar punctures due to surgery and reported no experience of subjective cognitive decline. However, as previously applied in the DDI cohort ^23^, here we employ an actuarial definition of cognitive normalcy and mild cognitive impairment based on neuropsychological test battery performance (see supplementary methods for details). Inclusion criteria are ages between 40 and 80 years and native language of Norwegian, Swedish, or Danish. The exclusion criteria are intellectual disability or other developmental disorders, brain trauma, stroke, dementia, severe psychiatric conditions, or severe somatic disease that might influence cognitive functions. The DDI protocol comprises extensive medical history, neurological and neuropsychological examinations, lumbar puncture, blood-draw and brain Magnetic resonance imaging (MRI).

### Cohort-2: Pre-symptomatic Evaluation of Experimental or Novel Treatments for Alzheimer’s Disease (PREVENT AD)

PREVENT AD is a longitudinal observational study consisting of 385 initially cognitively unimpaired older adults with a parental or multiple-sibling history of AD dementia^24^. Participants in the PREVENT-AD study are aged 60 years or older upon entry, or 55 years or older if within 15 years of their relative’s symptoms onset. All participants underwent Clinical Dementia Rating (CDR) and Montreal Cognitive assessment (MoCA) assessment upon enrolment^24^. These individuals underwent neuropsychological evaluation using Repeatable Battery for the Assessment of Neuropsychological Status (RBANS), MRI and blood draw for routine labs. A subsample of participants underwent Positron Emission Tomography (PET) scans of Aβ pathology.

### CSF and blood proteomics

In cohort-1 CSF Aβ1-42 and CSF Aβ1-40 concentrations were measured by the QuickPlex SQ 120 system from Meso Scale Discovery (MSD, MD, USA). The Aβ42/40 ratio was used to determine Aβ plaque pathology (cut-off ≤0.077)^25^. CSF samples included prior to October 2020 used commercial enzyme-linked immunosorbent assays (ELISAs) from Innotest, Fujirebio, Ghent, Belgium based on monoclonal antibodies to determine CSF phosphorylated tau (p-tau181) concentration. Due to a change in laboratory equipment, CSF samples included after October 2020 used Elecsys p-tau181 kits (n=421 were determined with Innotest (>65 pg/mL); n=10 determined with Elecsys (>19 pg/mL)^23^.

All the p-tau markers in cohort 1 and 2 were measured on the Simoa HD-X platform with one in thirty dilution factor in CSF and two fold factor in plasma. Plasma p-tau181 was measured according to the Karikari et al. method^7^, plasma p-tau231 by the published method by Ashton et al.^26^. Plasma UGOT p-tau217 Gonzalez-Ortiz et al. method^27^. Signal variations within and between analytical runs were assessed using three internal quality control samples at the beginning and the end of each run.

### Aβ-PET

In cohort-2 Aβ-[18F NAV4694] PET scans were performed at the McConnell Brain Imaging Centre at the Montreal Neurological Institute (MNI). Aβ-PET scans were obtained 40–70 minutes after injection (220 MBq). The images were reconstructed using a three-dimensional (3D) ordinary Poisson ordered subset expectation maximum ([OP-OSEM] algorithm with 10 iterations and 16 subsets^28^. The data was pre-processed by our in-house protocol that is available on (https://github.com/villeneuvelab/vlpp). Standardized uptake value ratios (SUVRs) were calculated for each region of the Desikan-Killaney atlas by dividing the tracer uptake in the cerebellar grey matter for Aβ-PET scans. A global amyloid index SUVR threshold of 1.27, equivalent to CL = 18, was selected for Aβ-PET positivity.

### Study design

Cohort-1 included cases and controls (n=431) according to the following criteria: 1) Cognitively normal (controls or SCD) with normal CSF Aβ42/40 ratio and normal p-tau181 biomarkers (CN A-/T-, n=169). 2) CN or MCI participants with pathological Aβ42/40 ratio, but normal p-tau181 (All A+/T-, n=50; CN=26; MCI=24). 3) pathological Aβ42/40 ratio and p-tau181 (All A+/T+, n=145; CN=40; MCI=105) and normal Aβ42/40 ratio but pathological and p-tau181 (All A-/T+, n=67; CN=34; MCI=33). Details of available markers for each group are detailed in table 1 & 2. A subsample had available glomerular filtration rate (GFR) as a kidney function test (n=335). Verbal episodic memory impairment is acknowledged as one of the earliest clinical features of AD.^29^ Thus, only delayed verbal memory recall was included in our analyses. Of the 431 included cases, n=418 (Aβ-, n=227; Aβ+, n=191) had available longitudinal CERAD word list verbal memory recall ^30^ assessments up to 9.67 years from baseline (Mean=3.55, SD=1.87, range =0.52 – 9.67). See table S1 for details. Cohort-2 included cases and controls (n=190) in whom amyloid status was determined using Aβ PET and were classified as Aβ- CN= 119; Aβ+ CN= 49; Aβ+ MCI= 21. See table S2 for details.

**Table 1.**
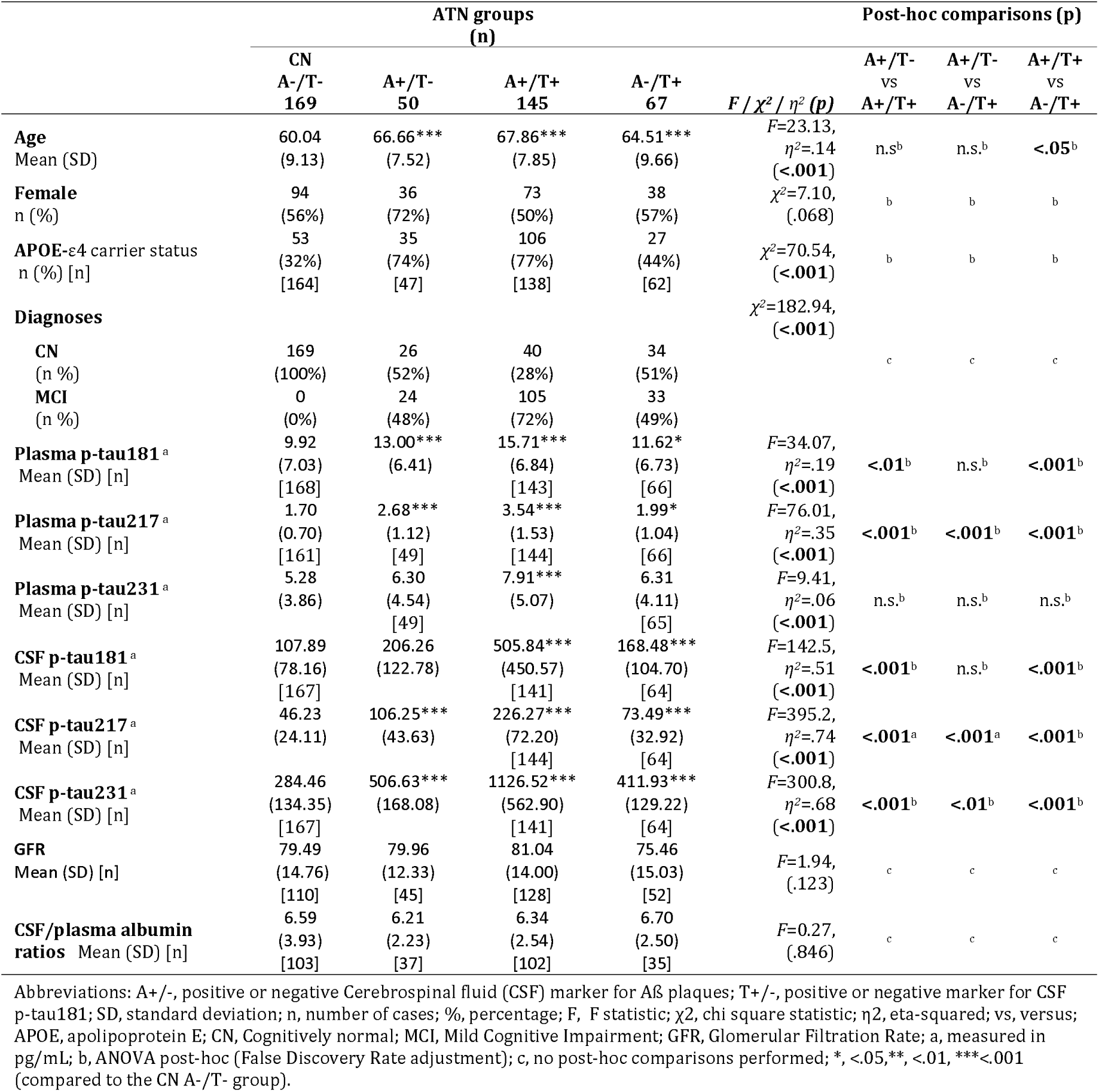
Between-group comparisons of demographics, *APOE-ε4* carrier status, diagnoses, plasma and CSF p-tau markers in cohort-1.

### Statistics

All analyses were performed in Rstudio (R version 4.3.2). Between-group differences in A/T groups in both cohorts (and CN/MCI Aβ-/Aβ+ for between cohort comparisons) for continuous variables (age, GFR and log-transformed CSF/Plasma p-tau biomarker concentrations) were assessed with one-way ANOVA and post-hoc comparisons performed with false discovery rate (FDR) adjustments. Categorical variables (sex, diagnoses, and *APOE* genotype) were assessed with chi-square tests. A sub analysis of the biomarker differences between CN and MCI cases within the pathological A/T groups were assessed with independent sample t-tests. Spearman’s rho correlations were performed between CSF and plasma p-tau epitopes in both the complete sample, and within A/T groups. Fisher z-transformation was used to compare the correlation coefficients. Relative mean change in both CSF and plasma biomarkers were computed for CN and MCI pathological A/T groups (B) with the mean biomarker concentrations for the CN A-/T- group (CN Aβ- for between cohort comparisons) as the reference 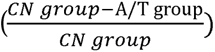. Receiver Operating Curve (ROC) analyses were performed for cognitive (CN A- vs CN A+ and MCI A+) and biological status (CN A-/T- vs A+/T- and A+/T+) and compared with Delonǵs test. Cut-offs for each model were generated using the Youden index, and NPVs and PPVs were computed accordingly. Linear Mixed Models were fitted to assess associations between baseline p-tau epitopes in CSF/Plasma and future memory decline (CERAD word list recall subtest ^30^) for A- (A-/T- & A+/T-) and A+ (A+/T- & A+/T+) separately. Spearman’s rho correlations between the plasma biomarkers and GFR were performed in the complete sample. See supplementary methods for additional details.

## Results

### Agreements between CSF and plasma p-tau biomarkers

In cohort-1, we observed a moderate correlation between CSF and plasma p-tau217 (rho=.65, p<.001, figure 1A), and significantly weaker correlations for p-tau181 (z= 3.83, p<.001, rho=.47, p<.001, figure 1B) and p-tau231 (z= 6.38, p<.001, rho=.31, p<.001, figure 1C). Split by A/T groups, p-tau217 correlations (figure 1D) were similar in both A+/T- (rho=.49, p<.001) and A+/T+ (rho=.48, p<.001) groups. Weaker correlations were seen in the CN A-/T- and A-/T+ groups (both rho=.24, p<.01; p=.056). For both p-tau181 (figure 1E) and p-tau231 (figure 1F), correlations were weaker in all groups as compared to p-tau217 (rhós between 0.14 and .28). Here, both p-tau181 and p-tau231 showed more robust correlations in the A+/T+ group (both rho=0.27, p<.001). Notably, the comparable performance of p-tau217 in A+/T- and A+/T+ groups highlights its ability to reliably capture CSF pathological changes, even in early stages of AD. In contrast, the weaker and more variable correlations for p-tau181 and p-tau231 suggest that these markers may be less sensitive to CSF-related biomarker abnormalities, limiting their utility in aligning plasma measurements with underlying pathology.

**Figure 1.**
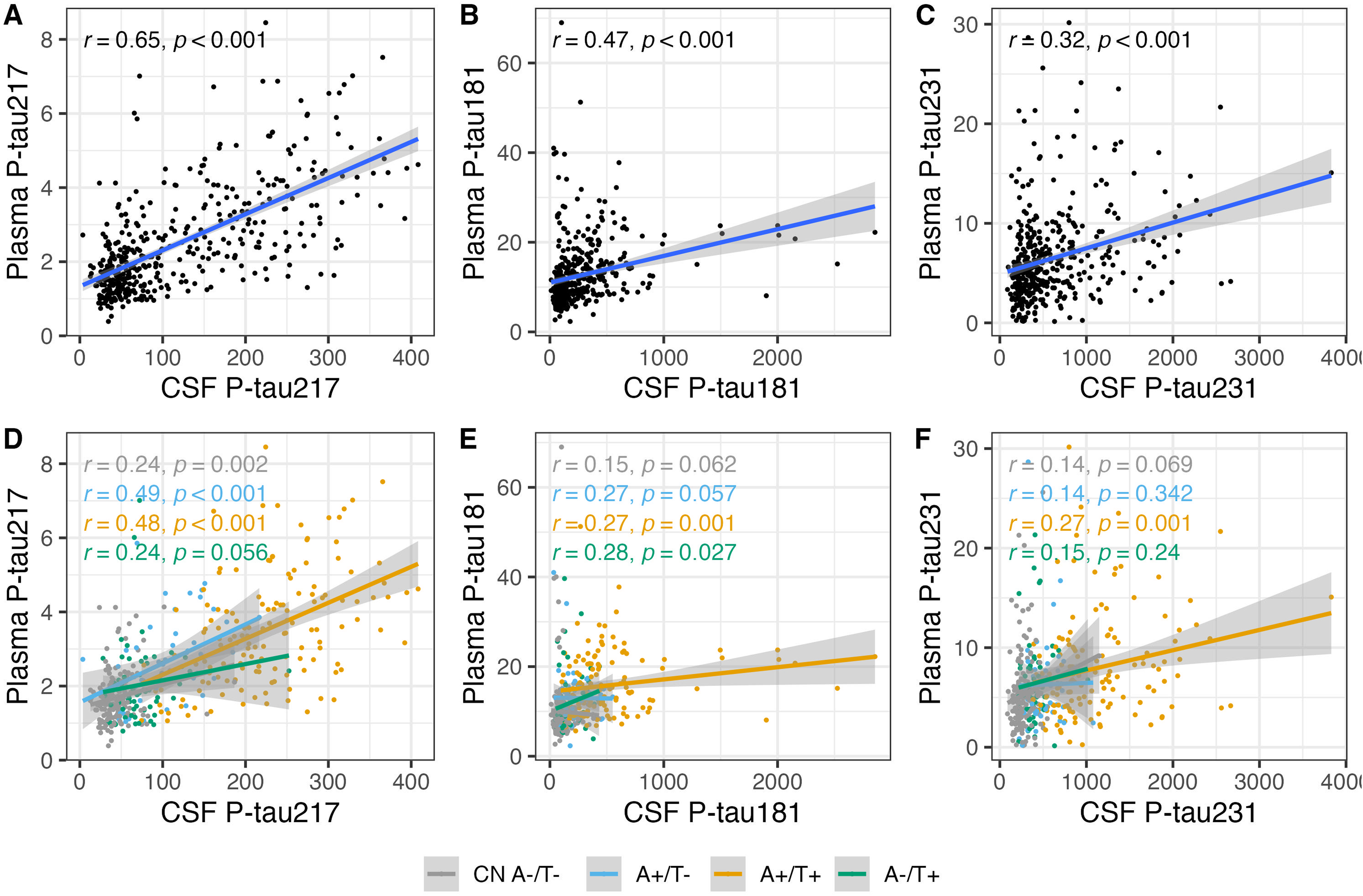
Agreements between CSF and plasma p-tau biomarkers in cohort-1. Scatterplots illustrating the spearman’s rho correlations between plasma and CSF p-tau markers **Figure 1A-C)** show the correlations of plasma p-tau217, p-tau181 and p-tau231 with their corresponding CSF markers. **Figure 1D-F)** show the CSF-plasma correlations of p-tau217, p-tau181 and p-tau231 in the different A/T groups.

### Diagnostic performance based on cognitive status versus biological status

Regardless of cognitive status, in cohort-1 plasma p-tau217 demonstrated the highest AUC to detect Aβ pathology (.850), followed by p-tau181 (.797) and p-tau231 (.661). However, only p-tau217 showed a significant increase in accuracy for MCI Aβ+ (.886) compared to CN Aβ+ (.786, p<.05) as well as A+/T- (.778) compared to A+/T+ (.876) (p<.05). See figure 2 and table S3A for details. Moreover, plasma p-tau217 demonstrated PPVs and NPVs above .800 for both MCI Aβ+ and A+/T+ versus A-/T- controls, but poor PPVs for CN Aβ+ and A+/T- (.656 & .458 respectively) See table S4A and figure S1 for details on all markers. In CSF, p-tau217 also showed the highest accuracy for Aβ pathology (.973) followed by p-tau231 (.961) and p-tau181 (.906). Here, all epitopes had higher accuracies for A+/T+ as compared to A+/T- (between p<.01 and p<.001). But only CSF p-tau231 differentiated between CN (.942) and MCI Aβ+ (.970) (p<.05). See figure S2, tables S3B and S4A for details.

**Figure 2.**
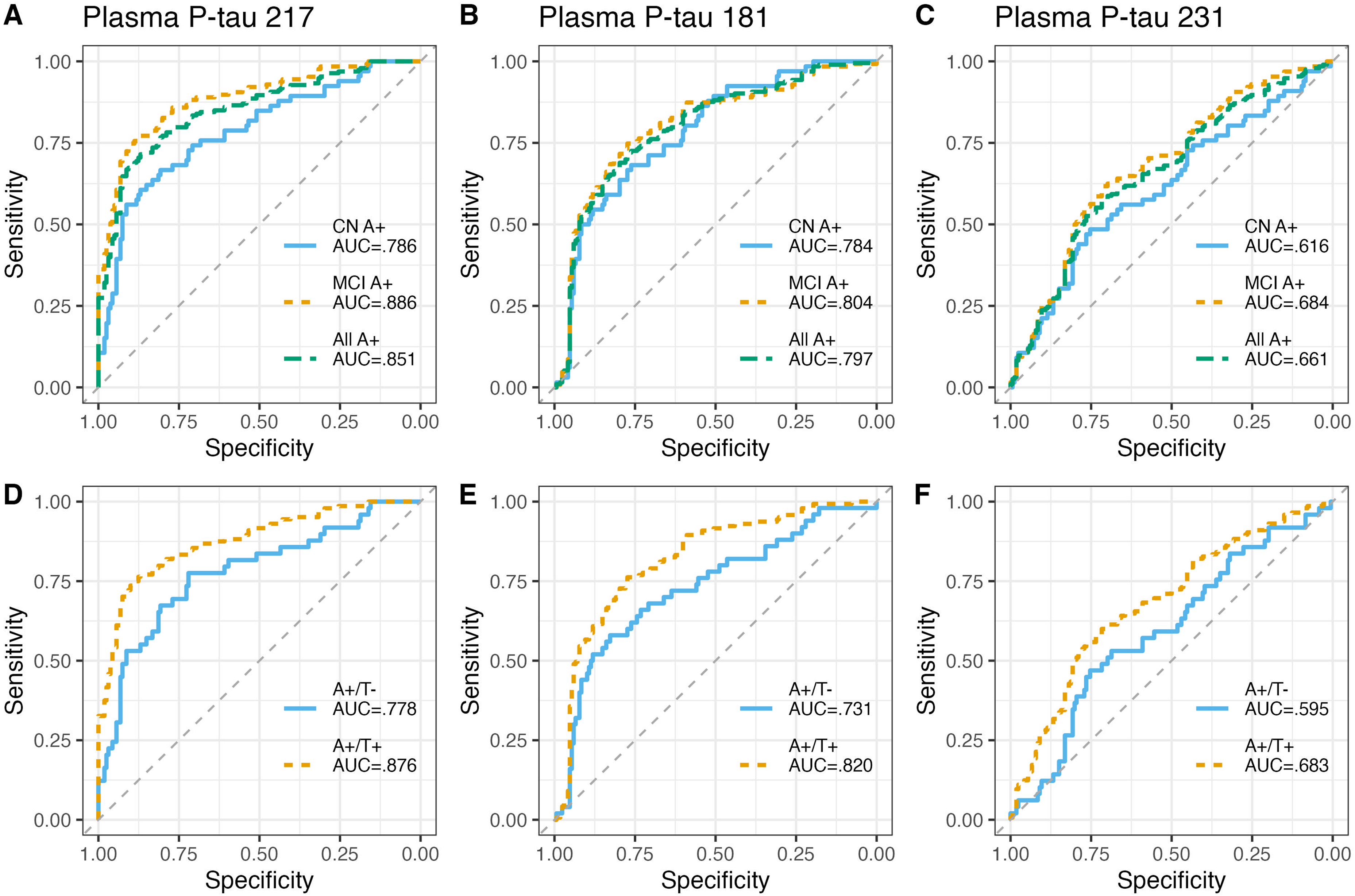
Diagnostic accuracy of plasma p-tau markers in cohort-1. Receiver Operating Characteristic (ROC) curves and corresponding areas under the curve (AUC) showing the discriminative ability of the different plasma p-tau biomarkers. **Figure 2A-C)** ROC curves and AUCs of plasma p-tau217, p-tau181 and p-tau231 identifying Aβ+ individuals based on their cognitive status. **Figure 2D-F)** ROC curves and AUCs of plasma p-tau217, p-tau181 and p-tau231 identifying Aβ+ individuals according to their A/T profile in CSF.

### Between-group differences in plasma and CSF p-tau and associations with memory decline

In CSF, p-tau217 and p-tau231 were higher in all pathological A/T groups (all p<.001), whereas p-tau181 was higher only in the A+/T+ and A-/T+ groups (both p<.001) See table 1 for details. When splitting by cognitive status, only plasma p-tau217 was higher in MCI A+/T+ compared to CN A+/T+ (p<.05), whereas only CSF p-tau217 and p-tau231 had higher concentrations in MCI A+/T+ compared to CN A+/T+ (p<.001) (see table S5). Relative mean change (with CN A-/T- as the reference) in biomarker concentrations corresponded to the between-group differences outlined above, notably demonstrating generally a larger relative mean change for both plasma and CSF p-tau217 in the Aβ+ groups as compared to the other p-tau epitopes, and also a higher relative mean change plasma p-tau 217 in MCI as compared to CN A+/T+ (see figure S3). Moreover, plasma p-tau217, but not p-tau181 or p-tau231, showed significant associations with both baseline (β=-0.32, p<.001) and future verbal memory decline (β=-0.05, p<.05) in Aβ+ but not in Aβ- cases (figure 3 and table S1). In CSF all p-tau epitopes associated with memory impairment and decline in Aβ+, however p-tau217 showed the strongest associations over time (β=-0.06, p<.05). P-tau181 was the only CSF p-tau marker to associate with future memory decline in the Aβ- cases (β=-0.04, p<.01) (see figure S4 and table S1 for details).

**Figure 3.**
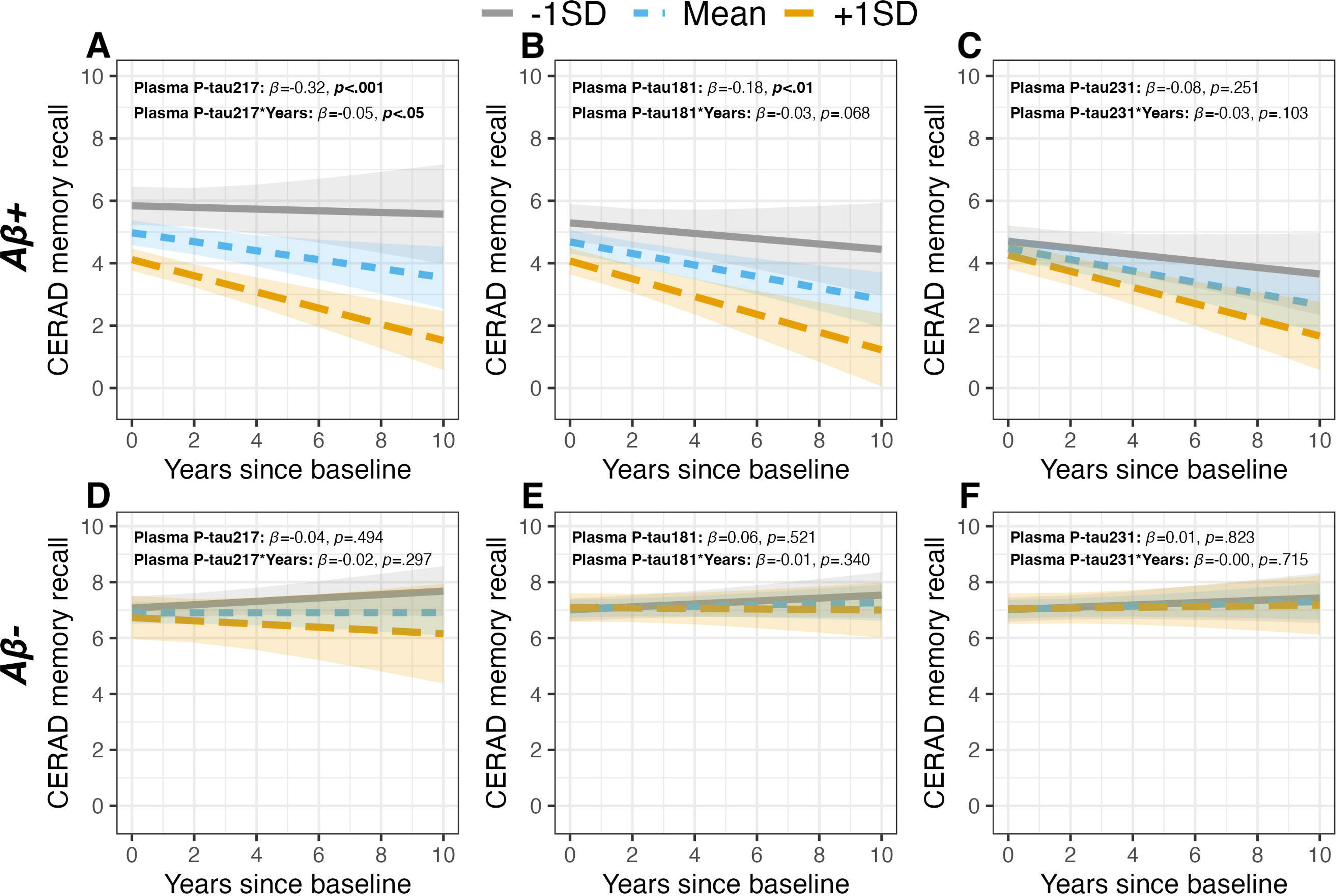
Baseline and longitudinal associations of plasma p-tau markers with the Consortium to Establish a Registry for Alzheimer’s Disease (CERAD) memory recall test in cohort-1. **Figure 3A-C)** show the baseline and longitudinal associations of plasma p-tau217, p-tau181 and p-tau231 with the CERAD memory recall test in Aβ+ individuals. **Figure 3D-F)** show the baseline and longitudinal associations of plasma p-tau217, p-tau181 and p-tau231 with the CERAD memory recall test in Aβ- individuals. The lines display associations between the biomarker at −1SD (grey), Mean (blue) and +1SD (orange) and the dependent variable at baseline and over time.

### Plasma p-tau biomarkers according to cognitive status

In cohort 1 and 2, we compared plasma p-tau217 concentrations between the CN Aβ+ (cohort 1: n=66; cohort 2: n=49) and MCI Aβ+ (cohort 1: n=127; cohort 2: n=21) groups to CN Aβ- (cohort 1: n=161; Cohort 2: n=118). In both cohorts, we found significantly higher p-tau217 concentrations in the MCI Aβ+ cases as compared to CN Aβ+ (both cohorts p<.001, see figure 4A&B). In cohort 1, we also compared plasma p-tau181 and p-tau231 between groups but found no difference in concentrations between CN Aβ+ and MCI Aβ+ cases (both n.s., see figure S5). Moreover, relative mean changes in p-tau217 for the Aβ+ groups (compared to CN Aβ-) were remarkably similar in both cohorts (figure 4C). Thus, replicating between independent cohorts that plasma p-tau217 is sensitive to cognitive severity in predementia AD regardless of the method used to determine Aβ positivity. Results of plasma p-tau181 and p-tau231 in cohort-2 have previously been reported^28^.

**Figure 4.**
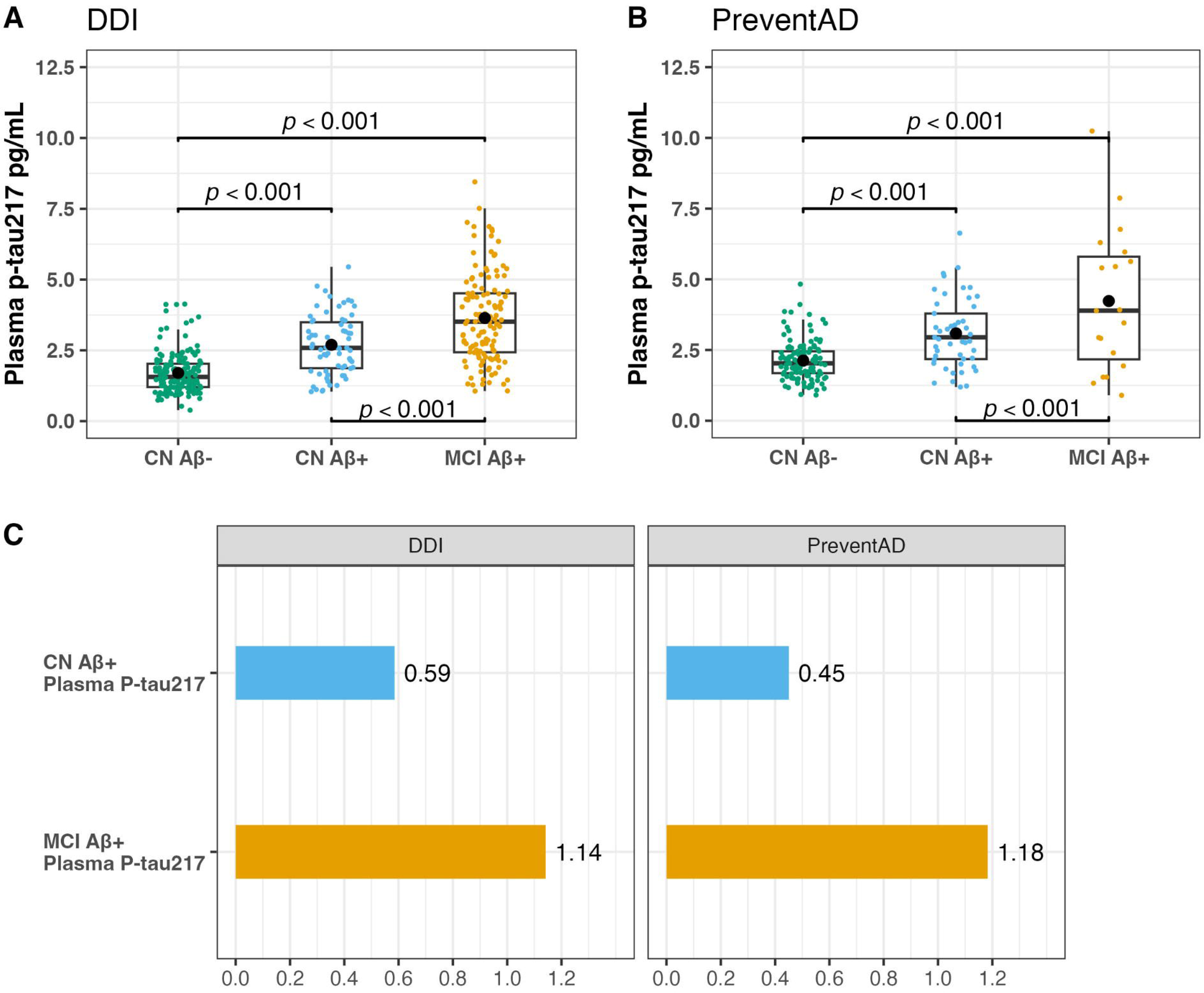
Plasma p-tau217 concentrations and relative mean change increases according to cognition in cohort 1 and 2. Figure 4A and **B)** Boxplots showing concentrations of plasma p-tau217 (pg/ml) in CN Aβ-, CN Aβ+ and MCI Aβ+ individuals in cohort 1 and 2. The brackets show statistically significant differences between the groups (FDR adjusted p-values). **Figure 4C)** The Bar graphs illustrate the relative mean change increases of plasma p-tau217 in Aβ+ CN and Aβ+ MCI participants compared with CN Aβ- in cohort 1 and cohort 2.

### Biomarker correlations with glomerular function

In cohort-1 plasma p-tau217 showed the weakest correlation with GFR (rho=-14, p<.05), followed by p-tau181 (rho=-17, p<.01) and p-tau231 (rho=-22, p<.001). As expected, no associations between GFR and CSF p-tau epitopes were found. No differences in GFR between the A/T groups were observed (Table 1).

## Discussion

While all p-tau markers in CSF demonstrated excellent diagnostic and prognostic accuracy, only plasma p-tau217 exhibited diagnostic and prognostic performance comparable to CSF p-tau markers. The superior performance of plasma p-tau217 over p-tau181 and p-tau231 at detecting early AD biochemical signatures and its sensitivity to capture cognitive changes might be attributed to its unique properties observed in-vitro models, such as promoting synaptic decline and the formation of tau-tau interactions at the expense of tau binding to microtubules^31^.

In cohort-1, when we divided Aβ+ participants according to their cognitive status (CN Aβ+ and MCI Aβ+) plasma p-tau217 showed significant elevation in the MCI Aβ+ group compared with CN Aβ+ group while plasma p-tau181 and p-tau231 did not show any difference between these groups. We replicated the plasma p-tau217 results in cohort-2, in which AD pathology was determined by Aβ-PET, where higher levels of plasma p-tau217 were associated to worse cognitive performance in Aβ+ participants. Moreover, relative mean change in biomarker concentrations compared to the control group were remarkably similar in the CN Aβ+ and MCI Aβ+ groups for both cohorts (Figure 4).

When dividing Aβ+ participants in cohort-1 according to their CSF profiles (A+/T- and A+/T+), we observed that in the A+/T- group none of the CSF or plasma biomarkers could differentiate between CN and MCI individuals. On the other hand, all the CSF p-tau markers and plasma p-tau217, but not p-tau181 or p-tau231, were capable of distinguishing between CN and MCI in the A+/T+ group. These findings suggest that while cognitive deterioration might impact the levels of plasma p-tau217 in Aβ+ individuals, it is likely that the joint pathology (A+/T+) is the main driver of significant increases in plasma p-tau217. It is important to note, however, that in our study, T+ was determined using clinically approved CSF p-tau181 assays, rather than the more appropriate CSF p-tau217 assay. Relatedly, we also demonstrate that relative mean changes for p-tau217 are greater in CSF than in plasma (Figure S3), with diagnostic concordance to CSF Aβ42/40-defined Aβ pathology being correspondingly higher in CSF. Furthermore, CSF p-tau217 outperforms CSF p-tau181 in this context^32^, underscoring its robustness as a marker of core AD pathology when measured in CSF compared to plasma. While it was recently suggested that plasma p-tau217 may show similar accuracies to clinically approved CSF based Aβ42/40 or p-tau181/Aβ42 diagnostics^33^, we propose that CSF p-tau217 likely represents a more appropriate benchmark for assessing the diagnostic performance of plasma p-tau biomarkers.

When comparing the performance of CSF versus plasma p-tau217 in cohort-1, we observe that even with less pronounced elevation in biomarker concentrations compared to controls, plasma p-tau217 was better able to identify Aβ+ participants. Furthermore, when we assessed the PPV and NPV of plasma p-tau markers, plasma p-tau217 showed a superior performance identifying true positive (TP) and true negative (TN) participants when compared with p-tau181 and p-tau231 (figure S1 and table S4A). However, all the plasma p-tau markers showed a poor performance identifying TP participants in the CN A+ and in the A+/T- groups. Our results suggest that the optimal diagnostic performance of plasma p-tau217, based on PPV and NPV, was achieved in the Aβ+ MCI participants and in those with an A+/T+ profile in CSF regardless of the cognitive status. These findings are particularly relevant for real-world settings, where relying solely on a single measurement of plasma p-tau217 might lead to misdiagnosis, if this is not taken in consideration. Lower PPVs for preclinical AD cases have been reported previously^13^ and are thought to result from the lower pre-test probability of amyloid pathology in cognitively normal individuals^34^. However, as we and others show^15,35^, higher concentrations of plasma p-tau217 correspond to degree of clinical impairment and future decline and may thus track pathological and clinical severity^36^. This is important, as not all CN A+ participants will develop MCI or dementia in their lifetimes^37^. Moreover, CN participants with higher concentrations of plasma p-tau217 may be at greater risk, and two cut-off approaches^38^ aimed at reducing the diagnostic mismatch with CSF or amyloid-PET may also serve to capture those with higher likelihood of clinical progression.

Interestingly, in Aβ+ cases, the association with future verbal memory decline was similar for plasma and CSF p-tau217. In contrast, CSF p-tau181 emerged as the only marker associated with memory decline in Aβ- cases (Figure 3 and Figure S4). Together, these findings underscore the specificity of p-tau217 as a core marker of AD pathology and highlight its prognostic value in both CSF and plasma. Furthermore, in cohort-1 we observed that plasma p-tau217 levels were less affected by kidney function when compared to p-tau181 and p-tau231, suggesting a higher robustness. These findings, in addition to the CSF-plasma correlations observed in cohort-1, might suggest that while in CSF the three p-tau markers are all sensitive for detecting AD-related pathology, in blood, potential peripheral contribution of p-tau181 and p-tau231 could affect the diagnostic and prognostic performance of these markers while plasma p-tau217 seems to be less affected by peripheral factors and a more accurate reflection of AD pathology.

### Conclusion

Levels of plasma p-tau217 align consistently with biological and clinical changes observed in AD and can provide valuable information about the course of the disease even in early stages, namely preclinical and prodromal AD. While p-tau181 and p-tau231 have been valuable in the context of AD research, studies like ours underscore the unique diagnostic and prognostic potential of plasma p-tau217 at capturing clinical severity. Moreover, our results address the potential limitations on the use of plasma p-tau217 very early in the clinical trajectory of AD while also acknowledging that, due to its minimally invasive nature and accessibility, plasma p-tau217 makes an excellent alternative for screening and routine clinical assessments when CSF analysis or PET are not available. Integrating plasma p-tau217 into clinical practice holds promise not only for improving AD diagnosis but also to facilitate early interventions in patients at risk of cognitive decline.

### Strengths and Limitations

The main strengths of our study include the inclusion of two independent cohorts with extensive neuroimaging and biomarker characterization of patients in preclinical and prodromal AD. The main limitation of this study was the lack of longitudinal blood sampling and the low racial and ethnical diversity of our population.

## Data Availability

Anonymized aggregated level data will be shared by request from a qualified academic investigator for the sole purpose of replicating procedures and results presented in the article, and as long as data transfer is in agreement with EU legislation on the general data protection regulation and decisions by the Ethical Review Boards in charge of each of the cohorts used for this study.

## Author Contributions

Kirsebom had full access to all the data from cohort 1 in the study and takes responsibility for the integrity of the data and the accuracy of the data analysis. Yakoub had full access to the data from cohort 2 and takes responsibility for the integrity of the data and the accuracy of the data analysis.

Concept and design: Gonzalez-Ortiz, Kirsebom, Fladby and Blennow

Acquisition, analysis, or interpretation of data: Gonzalez-Ortiz, Kirsebom, Yakoub, Gundersen and Gísladóttir.

Drafting of the manuscript: Gonzalez-Ortiz and Kirsebom.

Critical revision of the manuscript for important intellectual content: Gonzalez-Ortiz, Kirsebom, Pålhaugen, Waterloo, Selnes, Jarnholm, Rongve, Skogseth, Bråthen, Aarsland, Turton, Harrison, Zetterberg and Villeneuve.

Statistical analysis: Kirsebom and Yakoub.

Supervision: Fladby and Blennow.

## Conflict of interest

KB has served as a consultant and at advisory boards for Acumen, ALZPath, AriBio, BioArctic, Biogen, Eisai, Lilly, Moleac Pte. Ltd, Novartis, Ono Pharma, Prothena, Roche Diagnostics, and Siemens Healthineers; has served at data monitoring committees for Julius Clinical and Novartis; has given lectures, produced educational materials and participated in educational programs for AC Immune, Biogen, Celdara Medical, Eisai and Roche Diagnostics; and is a co-founder of Brain Biomarker Solutions in Gothenburg AB (BBS), which is a part of the GU Ventures Incubator Program, outside the work presented in this paper. HZ has served at scientific advisory boards and/or as a consultant for Abbvie, Acumen, Alector, Alzinova, ALZPath, Amylyx, Annexon, Apellis, Artery Therapeutics, AZTherapies, Cognito Therapeutics, CogRx, Denali, Eisai, Merry Life, Nervgen, Novo Nordisk, Optoceutics, Passage Bio, Pinteon Therapeutics, Prothena, Red Abbey Labs, reMYND, Roche, Samumed, Siemens Healthineers, Triplet Therapeutics, and Wave, has given lectures in symposia sponsored by Alzecure, Biogen, Cellectricon, Fujirebio, Lilly, Novo Nordisk, and Roche, and is a co-founder of Brain Biomarker Solutions in Gothenburg AB (BBS), which is a part of the GU Ventures Incubator Program (outside submitted work). BEK has served as a consultant for Biogen and advisory boards for Eisai and Eli Lilly. TF has served as a consultant and at the advisory boards for Biogen, Eisai, Novo Nordisk, Eli Lilly and Roche. RES has served on an advisory board for Eisai.

## Funding

The project was funded by the Norwegian Research Council, JPND/PMI-AD (NRC 311993). KB is supported by the Swedish Research Council (#2017-00915 and #2022-00732), the Swedish Alzheimer Foundation (#AF-930351, #AF-939721 and #AF-968270), Hjärnfonden, Sweden (#FO2017-0243 and #ALZ2022-0006), the Swedish state under the agreement between the Swedish government and the County Councils, the ALF-agreement (#ALFGBG-715986 and #ALFGBG-965240), the European Union Joint Program for Neurodegenerative Disorders (JPND2019-466-236), the Alzheimer’s Association 2021 Zenith Award (ZEN-21-848495), the Alzheimer’s Association 2022-2025 Grant (SG-23-1038904 QC), and the Kirsten and Freddy Johansen Foundation. HZ is a Wallenberg Scholar and a Distinguished Professor at the Swedish Research Council supported by grants from the Swedish Research Council (#2023-00356; #2022-01018 and #2019-02397), the European Union’s Horizon Europe research and innovation programme under grant agreement No 101053962, Swedish State Support for Clinical Research (#ALFGBG-71320), the Alzheimer Drug Discovery Foundation (ADDF), USA (#201809-2016862), the AD Strategic Fund and the Alzheimer’s Association (#ADSF-21-831376-C, #ADSF-21-831381-C, #ADSF-21-831377-C, and #ADSF-24-1284328-C), the Bluefield Project, Cure Alzheimer’s Fund, the Olav Thon Foundation, the Erling-Persson Family Foundation, Stiftelsen för Gamla Tjänarinnor, Hjärnfonden, Sweden (#FO2022-0270), the European Union’s Horizon 2020 research and innovation programme under the Marie Skłodowska-Curie grant agreement No 860197 (MIRIADE), the European Union Joint Programme – Neurodegenerative Disease Research (JPND2021-00694), the National Institute for Health and Care Research University College London Hospitals Biomedical Research Centre, and the UK Dementia Research Institute at UCL (UKDRI-1003). BEK was supported by a grant from Helse-Nord (HNF1540-20).

PREVENT-AD was launched in 2011 as a $13.5 million, 7-year public-private partnership using funds provided by McGill University, the Fonds de Recherche du Québec – Santé (FRQ-S), an unrestricted research grant from Pfizer Canada, the J.L. Levesque Foundation, the Lemaire Foundation, the Douglas Hospital Research Centre and Foundation, the Government of Canada, and the Canada Fund for Innovation. The PREVENT-AD was additionally supported by the Canadian Institute of Health Research (CIHR) (#438655), the Fonds de recherche du Québec en Santé (FRQS) and Brain Canada grants.

## Notes

### Competing Interest Statement

The authors have declared no competing interest.

### Funding Statement

Authors funding information is provided in the manuscript

### Summary of Updates

Updated manuscript after first round of reviews

