## Supplementary material 1 for "Impact of clinical severity on plasma p-tau performance in predementia Alzheimer’s disease"

16.- Department of Old Age Psychiatry. Institute of psychiatry, Psychology and Neuroscience King’s College London, London, UK.

17.- Bioventix Plc, 7 Romans Business Park, East Street, Farnham, Surrey GU9 7SX, UK

18.- Department of Neurodegenerative Disease, UCL Institute of Neurology, Queen Square, London, UK

19.- UK Dementia Research Institute at UCL, London, UK

20.- Hong Kong Center for Neurodegenerative Diseases, Clear Water Bay, Hong Kong, China

21.- Wisconsin Alzheimer’s Disease Research Center, University of Wisconsin School of Medicine and Public Health, University of Wisconsin-Madison, Madison, WI, USA

22.- Paris Brain Institute, ICM, Pitié-Salpêtrière Hospital, Sorbonne University, Paris, France

23.- Neurodegenerative Disorder Research Center, Division of Life Sciences and Medicine, and Department of Neurology, Institute on Aging and Brain Disorders, University of Science and Technology of China and First Affiliated Hospital of USTC, Hefei, P.R. China

*****Joint first authors

**^†^**Joint senior authors

**Correspondence:**

Fernando Gonzalez-Ortiz, Clinical Neurochemistry Lab House V3, floor 2 Mölndal Hospital

Street Address: Biskopsbogatan 27 SE-43180 Mölndal, Sweden.

Bjørn-Eivind Kirsebom, Neurology department, University Hospital of North Norway Hansine Hansens veg 67, 9019 Tromsø, Norway.

**Key words:** Predementia, CSF, Plasma, PET, Amyloid, Tau

**Supplementary methods**

*Applied algorithm for assessing cognitive normalcy and impairment.*

The present study included cases and controls from the Dementia Disease Initiation (DDI) cohort. The DDI cohort includes predementia cases with either SCD or MCI staged according to published criteria ^1,2^ and participants recruited as controls primarily from spouses of recruited patients, and secondarily from advertisements in local news media and also orthopaedic patients who had lumbar punctures due to surgery and reported no experience of subjective cognitive decline. However, for this and also for previous study^3^, we here employed an actuarial definition of cognitive normalcy and mild cognitive impairment based on neuropsychological test battery performance. This entails regarding all cases with at least one or more impaired cognitive scores as MCI, and cases with all scores normal as cognitively normal (CN), regardless of subjectively reported symptoms. This is supported by evidence showing that subjectively reported symptoms, in conjunction with objectively assessed impairment may not be a reliable criterion for MCI^4^. Here, we determined MCI when results were 1.5 SD below the normative mean within one or more cognitive domains. Our neuropsychological battery included the following domains/tests: 1) Delayed memory recall (Consortium to Establish a Registry for Alzheimer’s Disease (CERAD) word list delayed recall)^5^ , 2) executive function (Trail Making Test part B (TMT-B))^5^, 3) language/verbal fluency (Controlled Oral Word Association Test (COWAT))^6^ and 4) visuoperceptual ability (Visual Object and Space Perception Battery (VOSP) silhouettes)^7^. This procedure identified that n=9 (9.3%) of those recruited as controls had one or more impaired neuropsychological test scores and treated as MCI. Of these, none had normal A/T markers (n=2, A+/T-; n=3 A+/T+; n= 4, A-/T+). See the table below for a detailed overview of the cases within each A/T split by CN & MCI.

| Overview of cases recruited as controls, SCD and MCI in CN/MCI A/T groups. | | | | | | | |
| --- | --- | --- | --- | --- | --- | --- | --- |
|  | CN A-/T- | CN **A+**/T- | MCI **A+**/T- | CN **A+/T+** | MCI **A+/T+** | CN A-/**T+** | MCI A-/**T+** |
| Recruited as controls n (%) | 58 (34.3) | 8 (30.8) | 2 (8.3) | 8 (20) | 3 (2.9) | 14 (41.2) | 4 (12.1) |
| SCD  n (%) | 111 (65.7) | 18 (69.2) | 0 (0) | 32 (80) | 0 (0) | 20 (58.8) | 0 (0) |
| MCI  n (%) | 0 (0) | 0 (0) | 22 (91.7) | 0 (0) | 102 (97.1) | 0 (0) | 29 (87.9) |
| Abbreviations: CN, Cognitively Normal: MCI, Mild Conitive Impairment; SCD, Subjective Cognitive Decline; A+/-, pathological/normal CSF Aβ42/40 ratio; T+/-, pathological/normal CSF ptau181; n = number of cases. | | | | | | | |

Participants from the Prevent AD cohort are initially all cognitively unimpaired but are followed longitudinally. For the present study, all cases with available amyloid PET scans were evaluated. Over time, some PET amyloid positive cases developed MCI and their plasma samples were included in this study based on the closest timepoint after the participant’s MCI classification (Date plasma sample > Date MCI). If participants were cognitively unimpaired, the plasma timepoint closest to the PET scan (Positive or negative) was chosen. Overall, n = 190 participants were included in the study, n = 118 of whom were classified as cognitively unimpaired (CN) Aβ-PET negative, n = 50 CN Aβ-PET positive and n = 22 cognitively impaired Aβ-PET positive participants. MoCA and RBANS scores are calculated at baseline. Age and MMSE are calculated at time of PET-scan. The MMSE scores are only available for 178 participants. See table S2 for details.

*Supplementary statistics*

Analyses were performed in Rstudio (R version 4.3.2). For AUC, NPV, PPV and Delong´s tests, the “pROC” package was used^8^. All plots were generated using the “ggplot2”^9^ and “ggpubr”^10^ packages. Using the “lme4” and “lmerTest” packages^11,12^, Linear Mixed Models (LMMs) were fitted to assess associations between baseline p-tau epitopes in CSF/Plasma and future memory decline (CERAD word list recall subtest) for A- (A-/T- & A+/T-) and A+ (A+/T- & A+/T+) separately. Covariates included age at baseline, years of education and sex and were chosen due to their influence on CERAD delayed recall in a previous normative study^5^. A detailed account of covariate influence on the CERAD delayed recall scores for each model can be found in table S1. All models were fitted with random intercept for subject and random slope for time (Years since baseline). To allow for comparisons of relative strength between biomarkers and memory decline over time between models, all continuous variables, apart from years since baseline, were standardized (z-standardization). We used the “ggeffects”^13^ package to generate a data frame based on the LMM predictions (unstandardized continuous variables), and plotted the results using the “ggplot2” package^9^. Due to slight differences in CSF/plasma p-tau epitopes measured at baseline (see table 1 in the manuscript), number of individual cases in each model, as well as observations over time varied slightly. Between 224 and 227 subjects (between 556 to 572 observations over time) in the Aβ- group, and between 188 and 191 subjects (between 404 to 412 observations over time). Similarly, mean follow-up time varied slightly (Aβ-, 3.37 – 3.39 Years; Aβ+, 3.22 – 3.23 Years since baseline). Please see table S1 (excel document) for a detailed overview.

| **Supplementary table 2 (S2).** Demographics and biomarker differences between-group for the PreventAD cohort. | | | | | |
| --- | --- | --- | --- | --- | --- |
|  |  | Groups | | |  |
|  | Full sample  (N = 190) | CN **Aβ**-  (N = 118) | CN **Aβ+**  ( N =50) | MCI **Aβ+**  ( N = 22) | Post hoc comparison |
| Age, years | 67.80 (5.02) | 67.33 (4.92) | 67.94  (5.28) | 70.04 (4.55) | ns |
| Education, years | 15.60 (3.05) | 15.89 (3.06) | 15.24 (2.96) | 15.04  (3.17) | ns |
| Sex,F:M(%F) | 138 (73) | 80 (68) | 40 (80) | 18 (82) | ns |
| *APOE* carriers, n(%) | 72 (37) | 32 (27) | 28 (56) | 12 (54) | < 0.001 ^a^  < 0.05 ^b^ |
| Global amyloid, SUVR | 1.31 (0.29) | 1.15 (0.06) | 1.52 (0.30) | 1.68 (0.40) | < 0.001 ^a,b,c^ |
| plasma p-tau217(pg/ml) | 2.73 (1.63) | 2.15 (0.70) | 3.23 (1.53) | 4.74 (3.03) | < 0.001 ^a,b,c^ |
| MoCA (/30) | 28.30 (1.50) | 28.15 (1.47) | 28.74 (1.26) | 28.00 (1.95) | ns |
| RBANS at baseline | 104.87(10.24) | 104.45 (9.10) | 103.62 (9.12) | 100.71 (8.40) | ns |
| RBANS at last follow-up visit | 103.29 (10.62) | 104.35 (10.50) | 104.49 (8.61) | 94.62 (11.94) | <0.001^b^  <0.001^c^ |
| MMSE (/30) | 28.93(1.20) | 29.06 (1.08) | 28.96  (1.23) | 28.15  (1.56) | <0.001 ^b^  <0.05 ^c^ |
| *Abbreviations:* ^a^ difference between CN Aβ- and CN Aβ+; ^b^ differences between CN Aβ- and MCI Aβ+; ^c^ differences between CN Aβ+ and MCI Aβ+ individuals. Notes: three participants did not have p-tau217 values, one participant had missing MoCA score. F = female  *APOE* = apolipoprotein-E genotype; SUVR = Standardized Uptake Value Ratio; MoCA = Montreal Cognitive Assessment;  RBANS = Repeatable Battery for the Assessment of Neuropsychological status; ns = not significant.  *Notes:* One-way analysis of variance (ANOVA) with Tukey post hoc was used for continuous variables, and fisher tests for categorical variables. | | | | | |

| **Supplementary table 3A (S3A).** ROC analyses of plasma P-tau markers in Dementia Disease Initiation (Cohort 1) | | | | | | | | |
| --- | --- | --- | --- | --- | --- | --- | --- | --- |
|  | **Marker** | **Standard of truth** | **AUC  (95 % CI)** | **Cases/ controls** | **Spec.** | **Sens.** | **Cut-off** | **Delong´s test (p)** |
| A/T | P-tau 217 | A-/T- vs **A+**/T- | .778 (.699-.856) | 49/161 | .720 | .776 | ≥1.90 | ^c^ |
|  | P-tau 181 |  | .731 (.646-.815) | 50/168 | .827 | .580 | ≥11.89 | ^c^ |
|  | P-tau 231 |  | .595 (.505-.686) | 49/166 | .753 | .469 | ≥6.14 | ^c^ |
|  | P-tau 217 | A-/T- vs **A+**/**T+** | .876 (.837-.915) | 144/161 | .869 | .771 | ≥2.36 | -2.17 (**<.05**)^a^ |
|  | P-tau 181 |  | .820 (.773-.868) | 143/168 | .774 | .762 | ≥10.76 | -1.82 (n.s.)^a^ |
|  | P-tau 231 |  | .683 (.624-.743) | 145/166 | .789 | .531 | ≥6.57 | -1.59 (n.s.)^a^ |
| CN / MCI **A+** | P-tau 217 | CN A- vs  CN **A+** | .786 (.717-.854) | 66/161 | .717 | .854 | ≥2.35 | ^c^ |
|  | P-tau 181 |  | .784 (.720-.847) | 66/168 | .762 | .682 | ≥10.58 | ^c^ |
|  | P-tau 231 |  | .616 (.533-.698) | 66/166 | .753 | .485 | ≥6.14 | ^c^ |
|  | P-tau 217 | CN A- vs  MCI **A+** | .886 (.846-.925) | 127/161 | .901 | .748 | ≥2.44 | -2.47 (**<.05**)^b^ |
|  | P-tau 181 |  | .804 (.752-.857) | 127/168 | .774 | .748 | ≥10.76 | -0.49 (n.s.)^b^ |
|  | P-tau 231 |  | .684 (.602-.727) | 128/166 | .717 | .617 | ≥5.76 | -1.31 (n.s.)^b^ |
| All **A+** | P-tau 217 | CN A-/T- vs All **A+** | .851 (.812-.891) | 193/161 | .870 | .715 | ≥2.36 | ^c^ |
|  | P-tau 181 |  | .797 (.750-.844) | 193/168 | .774 | .720 | ≥10.76 | ^c^ |
|  | P-tau 231 |  | .661 (.605-.717) | 194/166 | .765 | .526 | ≥6.30 | ^c^ |
| Abbreviations*:* CSF= Cerebrospinal fluid; AUC = Area Under curve; CI= confidence interval. ^a^=Delong´s test CN vs MCI A+; ^b^=Delong´s test A+/T- vs A+/T+; ^c^=No Delong´s test performed. Please note that A+ and T+ were determined by CSF Mesoscale Discovery Aβ42/40 ratio and CSF innotest P-tau 181. In all models, the CN A-/T- cases are controls. Cut-off determined with the Youden index. | | | | | | | | |

| **Supplementary table 3B (S3B).** ROC analyses of CSF P-tau markers in Dementia Disease Initiation (Cohort 1) | | | | | | | |  |
| --- | --- | --- | --- | --- | --- | --- | --- | --- |
|  | **Marker** | **Standard of truth** | **AUC  (95 % CI)** | **Cases/ controls** | **Spec.** | **Sens.** | **Cut-off** | **Delong´s test (p)** |
| A/T | P-tau 217 | A-/T- vs **A+**/T- | .912 (.860-.961) | 50/169 | .893 | .820 | ≥66.38 | ^c^ |
|  | P-tau 181 |  | .794 (.729-.860) | 50/167 | .647 | .860 | ≥92.24 | ^c^ |
|  | P-tau 231 |  | .885 (.834-.935) | 50/167 | .850 | .800 | ≥371.09 | ^c^ |
|  | P-tau 217 | A-/T- vs **A+**/**T+** | .994 (.989-.999) | 144/169 | .964 | .986 | ≥86.81 | -3.23 (**<.01**)^a^ |
|  | P-tau 181 |  | .946 (.924-.968) | 141/167 | .844 | .879 | ≥170.63 | -4.32 (**<.001**)^a^ |
|  | P-tau 231 |  | .988 (.977-.998) | 141/167 | .964 | .950 | ≥488.86 | -3.89 (**<.001**)^a^ |
| CN / MCI **A+** | P-tau 217 | CN A- vs  CN **A+** | .957 (.932-.981) | 66/169 | .932 | .981 | ≥77.11 | ^c^ |
|  | P-tau 181 |  | .872 (.823-.872) | 66/167 | .683 | .909 | ≥104.58 | ^c^ |
|  | P-tau 231 |  | .934 (.900-.969) | 66/167 | .850 | .909 | ≥371.09 | ^c^ |
|  | P-tau 217 | CN A- vs  MCI **A+** | .981 (.964-.999) | 128/169 | .965 | .953 | ≥85.34 | -1.58 (n.s.)^b^ |
|  | P-tau 181 |  | .924 (.896-.952) | 125/167 | .814 | .864 | ≥160.76 | -1.83 (n.s.)^b^ |
|  | P-tau 231 |  | .975 (.958-.991) | 125/167 | .976 | .896 | ≥505.13 | -2.08 (**<.05)**^b^ |
| All **A+** | P-tau 217 | CN A-/T- vs All **A+** | .973 (.957-.989) | 194/169 | .964 | .907 | ≥83.54 | ^c^ |
|  | P-tau 181 |  | .906 (.877-.934) | 191/167 | .844 | .785 | ≥170.63 | ^c^ |
|  | P-tau 231 |  | .961 (.942-.979) | 191/167 | .976 | .827 | ≥505.13 | ^c^ |
| Abbreviations*:* CSF= Cerebrospinal fluid; AUC = Area Under curve; CI= confidence interval. ^a^=Delong´s test CN vs MCI A+; ^b^=Delong´s test A+/T- vs A+/T+; ^c^=No Delong´s test performed. Please note that A+ and T+ were determined by CSF Mesoscale Discovery Aβ42/40 ratio and CSF innotest P-tau 181. In all models, the CN A-/T- cases are controls. Cut-off determined with the Youden index. | | | | | | | |  |

| **Supplementary table 4A (S4A).** Positive and negative predictive values for Plasma p-tau markers in Dementia Disease Initiation (Cohort 1) | | | | |
| --- | --- | --- | --- | --- |
| **Versus** | **Plasma marker** | **CSF negative** | **CSF positive** | **NPV PPV** |
| **CN A- vs  CN A+** | **P-tau217** Negative (%) | 140 (84.3) TN | 26 (15.7) FN | .843 |
|  | **P-tau217** Positive (%) | 21 (34.4) FP | 40 (65.6) TP | .656 |
| **CN A- vs MCI A+** | **P-tau217** Negative (%) | 145 (81.9) TN | 32 (18.1) FN | .819 |
|  | **P-tau217** Positive (%) | 16 (14.4) FP | 95 (85.6) TP | .856 |
| **All A- vs  All A+** | **P-tau217** Negative (%) | 140 (71.8) TN | 55 (28.2) FN | .718 |
|  | **P-tau217** Positive (%) | 21 (13.2) FP | 138 (86.8) TP | .868 |
| **A-/T- vs A+/T-** | **P-tau217** Negative (%) | 116 (91.3) TN | 11 (8.7) FN | .913 |
|  | **P-tau217** Positive (%) | 45 (54.2) FP | 38 (45.8) TP | .458 |
| **A-/T- vs A+/T+** | **P-tau217** Negative (%) | 140 (80.9) | 33 (19.1) | .809 |
|  | **P-tau217** Positive (%) | 21 (15.9) | 111 (84.1) | .841 |
| **CN A- vs  CN A+** | **P-tau181** Negative (%) | 128 (85.9) TN | 21 (14.1) FN | .859 |
|  | **P-tau181** Positive (%) | 40 (47.1) FP | 45 (52.9) TP | .529 |
| **CN A- vs MCI A+** | **P-tau181** Negative (%) | 130 (80.2) TN | 32 (19.8) FN | .802 |
|  | **P-tau181** Positive (%) | 38 (28.6) FP | 95 (71.4) TP | .714 |
| **CN A- vs  CN A+** | **P-tau181** Negative (%) | 130 (70.7) TN | 54 (29.3) FN | .707 |
|  | **P-tau181** Positive (%) | 38 (21.5) FP | 139 (78.5) TP | .785 |
| **A-/T- vs A+/T-** | **P-tau181** Negative (%) | 139 (86.9) TN | 21 (13.1) FN | .869 |
|  | **P-tau181** Positive (%) | 29 (50.0) FP | 29 (50.0) TP | .500 |
| **A-/T- vs A+/T+** | **P-tau181** Negative (%) | 130 (79.3) | 34 (20.7) | .793 |
|  | **P-tau181** Positive (%) | 38 (25.9) | 109 (74.1) | .741 |
| **CN A- vs  CN A+** | **P-tau231** Negative (%) | 125 (78.6) TN | 34 (21.4) FN | .786 |
|  | **P-tau231** Positive (%) | 41 (56.2) FP | 32 (43.8) TP | .438 |
| **CN A- vs MCI A+** | **P-tau231** Negative (%) | 119 (70.8) TN | 49 (29.2) FN | .708 |
|  | **P-tau231** Positive (%) | 47 (37.3) FP | 79 (62.7) TP | .627 |
| **All A- vs  All A+** | **P-tau231** Negative (%) | 127 (58.0) TN | 92 (42.0) FN | .580 |
|  | **P-tau231** Positive (%) | 39 (27.7) FP | 102 (72.3) TP | .723 |
| **A-/T- vs A+/T-** | **P-tau231** Negative (%) | 125 (82.8) TN | 26 (17.2) FN | .828 |
|  | **P-tau231** Positive (%) | 41 (64.1) FP | 23 (35.9) TP | .359 |
| **A-/T- vs A+/T+** | **P-tau231** Negative (%) | 131 (65.8) | 68 (34.2) | .658 |
|  | **P-tau231** Positive (%) | 35 (31.2) | 77 (68.8) | .688 |
| Abbreviations*:* CSF= Cerebrospinal fluid; NPV = Negative Predictive Value; PPV = Positive Predictive Value; TP = True Positive; TN = True Negative; TN = True Negative; FN = False Negative. Please note that A+ and T+ were determined by CSF Mesoscale Discovery Aβ42/40 ratio and CSF innotest P-tau 181. In all models, the CN A-/T- cases are controls. Cut-off applied according to the Youden index for each model (see table 1A). | | | | |

| **Supplementary table 4B (S4B).** Positive and negative predictive values for CSF p-tau markers in Dementia Disease Initiation (Cohort 1) | | | | |
| --- | --- | --- | --- | --- |
| **Versus** | **CSF marker** | **CSF negative** | **CSF positive** | **NPV PPV** |
| **CN A- vs  CN A+** | **P-tau217** Negative (%) | 160 (94.1) TN | 10 (5.9) FN | .941 |
|  | **P-tau217** Positive (%) | 9 (13.8) FP | 56 (86.2) TP | .862 |
| **CN A- vs MCI A+** | **P-tau217** Negative (%) | 163 (96.4) TN | 6 (3.6) FN | .964 |
|  | **P-tau217** Positive (%) | 6 (4.7) FP | 122 (95.3) TP | .953 |
| **All A- vs  All A+** | **P-tau217** Negative (%) | 163 (90.1) TN | 18 (9.9) FN | .901 |
|  | **P-tau217** Positive (%) | 6 (3.3) FP | 176 (96.7) TP | .967 |
| **A-/T- vs A+/T-** | **P-tau217** Negative (%) | 151 (94.4) TN | 9 (5.62) FN | .944 |
|  | **P-tau217** Positive (%) | 18 (30.3) FP | 41 (69.5) TP | .695 |
| **A-/T- vs A+/T+** | **P-tau217** Negative (%) | 163 (98.8) | 2 (1.2) | .988 |
|  | **P-tau217** Positive (%) | 6 (4.1) | 142 (95.9) | .959 |
| **CN A- vs  CN A+** | **P-tau181** Negative (%) | 114 (95.0) TN | 6 (5.0) FN | .950 |
|  | **P-tau181** Positive (%) | 53 (46.9) FP | 60 (53.1) TP | .531 |
| **CN A- vs MCI A+** | **P-tau181** Negative (%) | 136 (88.9) TN | 17 (11.1) FN | .889 |
|  | **P-tau181** Positive (%) | 31 (22.3) FP | 108 (77.0) TP | .777 |
| **All A- vs  All A+** | **P-tau181** Negative (%) | 141 (77.5) TN | 41 (22.5) FN | .775 |
|  | **P-tau181** Positive (%) | 26 (14.8) FP | 150 (85.2) TP | .852 |
| **A-/T- vs A+/T-** | **P-tau181** Negative (%) | 108 (93.9) TN | 7 (6.1) FN | .939 |
|  | **P-tau181** Positive (%) | 59 (57.8) FP | 43 (42.2) TP | .422 |
| **A-/T- vs A+/T+** | **P-tau181** Negative (%) | 141 (89.2) TN | 17 (10.8) FN | .892 |
|  | **P-tau181** Positive (%) | 26 (17.3) FP | 124 (82.7) TP | .827 |
| **CN A- vs  CN A+** | **P-tau231** Negative (%) | 142 (95.9) TN | 6 (4.1) FN | .959 |
|  | **P-tau231** Positive (%) | 25 (29.4) FP | 60 (70.6) TP | .706 |
| **CN A- vs MCI A+** | **P-tau231** Negative (%) | 163 (92.6) TN | 13 (7.4) FN | .926 |
|  | **P-tau231** Positive (%) | 4 (3.4) FP | 112 (96.6) TP | .966 |
| **All A- vs  All A+** | **P-tau231** Negative (%) | 163 (83.2) TN | 33 (16.8) FN | .832 |
|  | **P-tau231** Positive (%) | 4 (2.5) FP | 158 (97.5) TP | .975 |
| **A-/T- vs A+/T-** | **P-tau231** Negative (%) | 142 (93.4) TN | 10 (6.6) FN | .934 |
|  | **P-tau231** Positive (%) | 25 (38.5) FP | 40 (61.5) TP | .615 |
| **A-/T- vs A+/T+** | **P-tau231** Negative (%) | 161 (95.8) | 7 (4.2) | .958 |
|  | **P-tau231** Positive (%) | 6 (4.3) | 134 (95.7) | .957 |
| *Abbreviations:* CSF= Cerebrospinal fluid; NPV = Negative Predictive Value; PPV = Positive Predictive Value; TP = True Positive; TN = True Negative; TN = True Negative; FN = False Negative. Please note that A+ and T+ were determined by CSF Mesoscale Discovery Aβ42/40 ratio and CSF innotest P-tau 181. In all models, the CN A-/T- cases are controls. Cut-off applied according to the Youden index for each model (see table 1B). | | | | |

| **Supplementary Table 5 (S5).** Between-group differences between CN and MCI within the pathological A/T groups split for p-tau epitopes and BD-tau in CSF and plasma. | | | | | | | | |
| --- | --- | --- | --- | --- | --- | --- | --- | --- |
|  | **A/T groups by cognitive status (n)** | | | | | | | |
|  | **CN A+/T-**  **26** | **MCI A+/T-**  **24** |  | **CN A+/T+**  **40** | **MCI A+/T+ 105** |  | **CN A-/T+ 34** | **MCI A-/T+ 33** |
| **Plasma p-tau181^a^** Mean (SD) [n] | 13.24 (6.63) | 12.73 (6.30) |  | 14.50 (5.72) | 16.19 (7.20) [103] |  | 11.96 (6.16) [33] | 11.28 (7.33) |
| **Plasma p-tau217^a^** Mean (SD) [n] | 2.42 (0.95) | 2.97 (1.25) [23] |  | **2.87 (1.09)** | **3.79** (1.60) [104]** |  | 1.99 (0.92) | 2.00 (1.17) [32] |
| **Plasma p-tau231^a^** Mean (SD) [n] | 5.54 (3.57) | 7.15 (5.39) [23] |  | 7.92 (5.33) | 7.91 (5.00) |  | 6.58  (4.01) [32] | 6.05 (4.26) |
| **CSF p-tau181^a^** Mean (SD) [n] | 218.23 (130.37) | 193.30 (115.33) |  | 413.75 (270.91) | 542.31 (500.87) [101] |  | 181.81 (103.56) [33] | 154.30 (105.71) [31] |
| **CSF p-tau217^a^** Mean (SD) [n] | 100.70 (46.90) | 112.26 (39.91) |  | **196.07 (70.95)** | **237.89*** (69.57) [104]** |  | 76.24 (40.76) [33] | 70.56 (22.05) [31] |
| **CSF p-tau231^a^** Mean (SD) [n] | 512.38 (195.36) | 500.40 (136.45) |  | **891.94 (477.55)** | **1,219.43*** (569.15) [101]** |  | 422.66 (157.67) [33] | 400.51 (91.02) [31] |
| *Abbreviations:* A+/-, positive or negative CSF marker for Aß plaques; T+/-, positive or negative marker for CSF p-tau181; CN, Cognitively normal; MCI, Mild Cognitive Impairment; SD, standard deviation; n, number of cases; ^a^, measured in pg/mL; *, <.05,**, <.01, ***<.001 (between CN and MCI within each A/T group) | | | | | | | | |

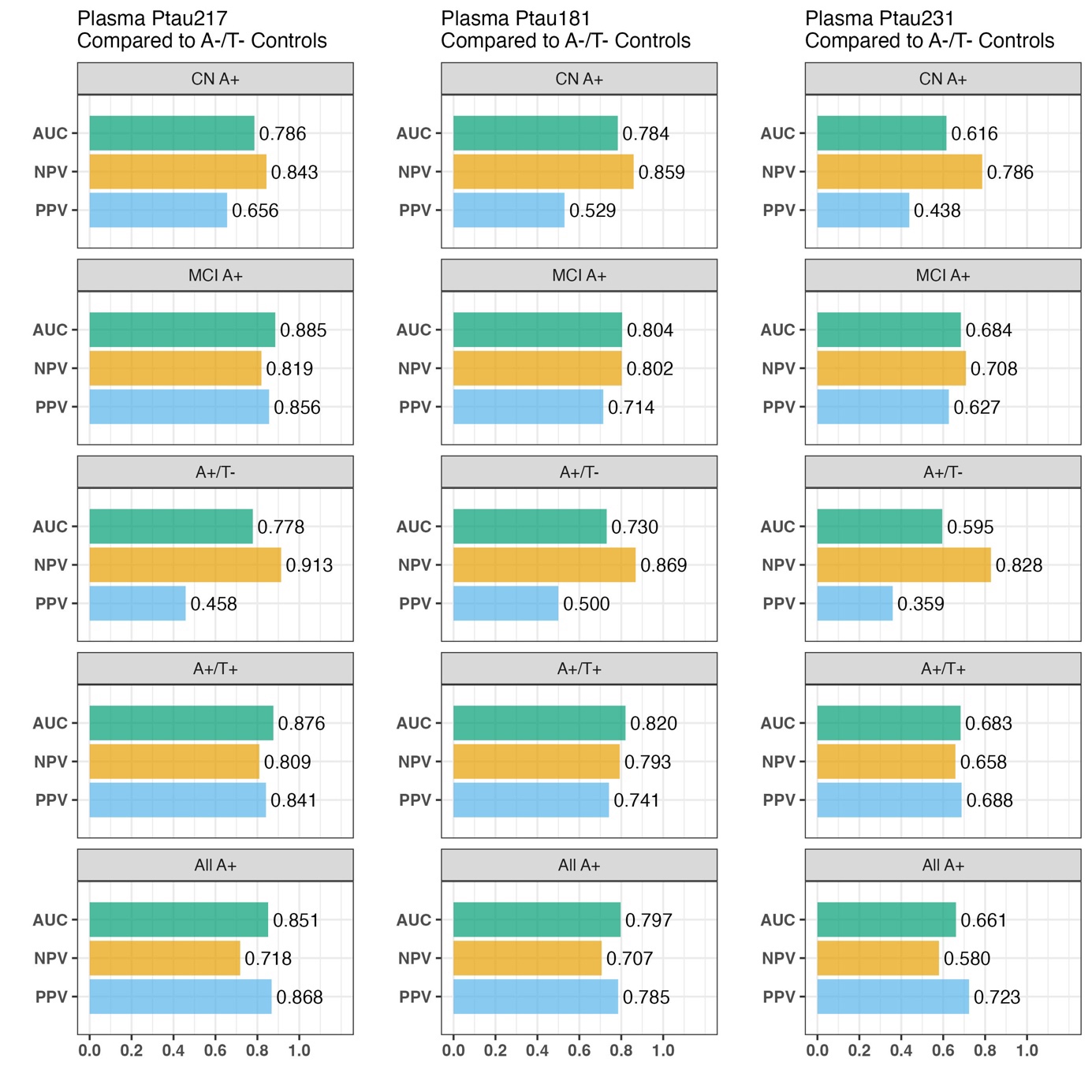

**Supplementary figure 1 (S1). Area Under Curve (AUC), Positive and negative predictive values (PPV, NPV) for the different Reciever Operating Curve models on cohort 1.** Shows a comparison of AUCs, PPVs and NPVs between the different ROC models performed in Dementia Disease Initation (cohort 1).

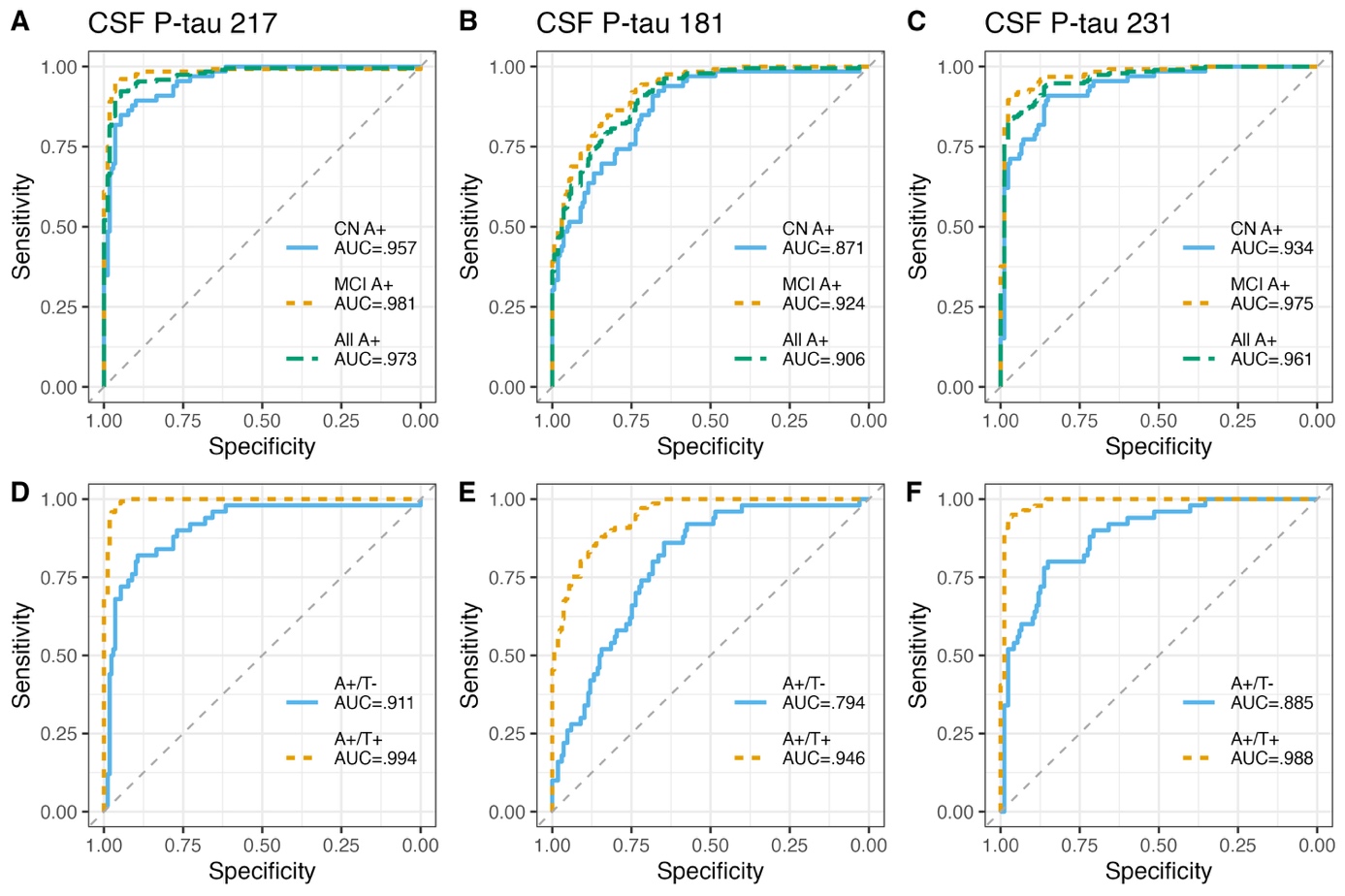

**Supplementary figure 2 (S2). Diagnostic accuracy of CSF p-tau markers in cohort-1.** Receiver Operating Characteristic (ROC) curves and corresponding areas under the curve (AUC) showing the discriminative ability of the different CSF p-tau biomarkers. **Figure S2A-C)** ROC curves and AUCs of CSF p-tau217, p-tau181 and p-tau231 identifying Aβ+ individuals based on their cognitive status. **Figure S2D-F)** ROC curves and AUCs of CSF p-tau217, p-tau181 and p-tau231 identifying Aβ+ individuals according to their A/T profile in CSF.

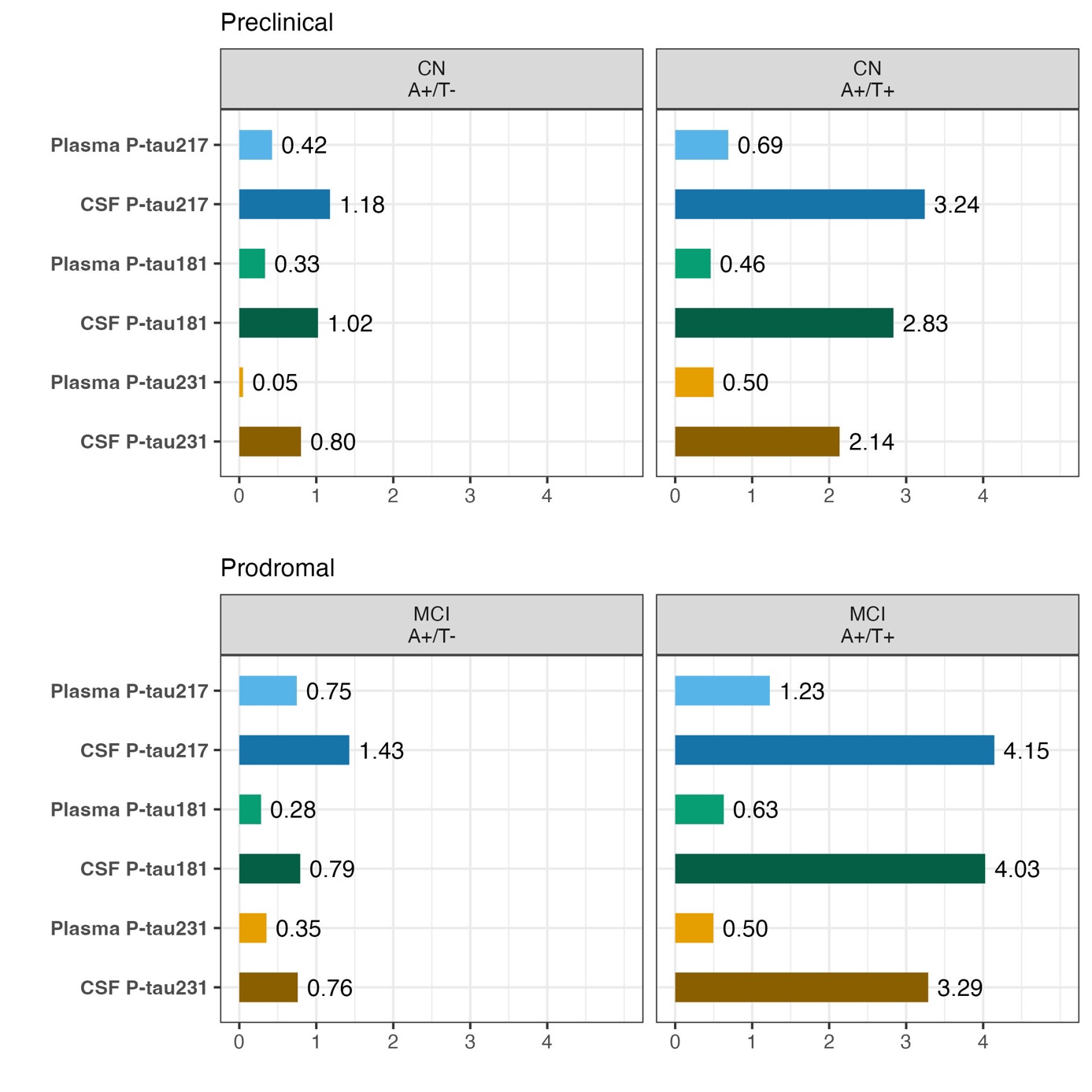

**Supplementary figure 3 (S3). Relative mean change increase in preclinical and prodromal cases in cohort 1.** This figure shows the relative mean change increase relative to cognitively normal (CN) A-/T- controls for the different p-tau epitopes measured in both plasma and CSF in cohort 1. The top figures shows cases with preclinical (CN A+), and prodromal (Mild Cognitive Impairment (MCI) A+), both split by T status.

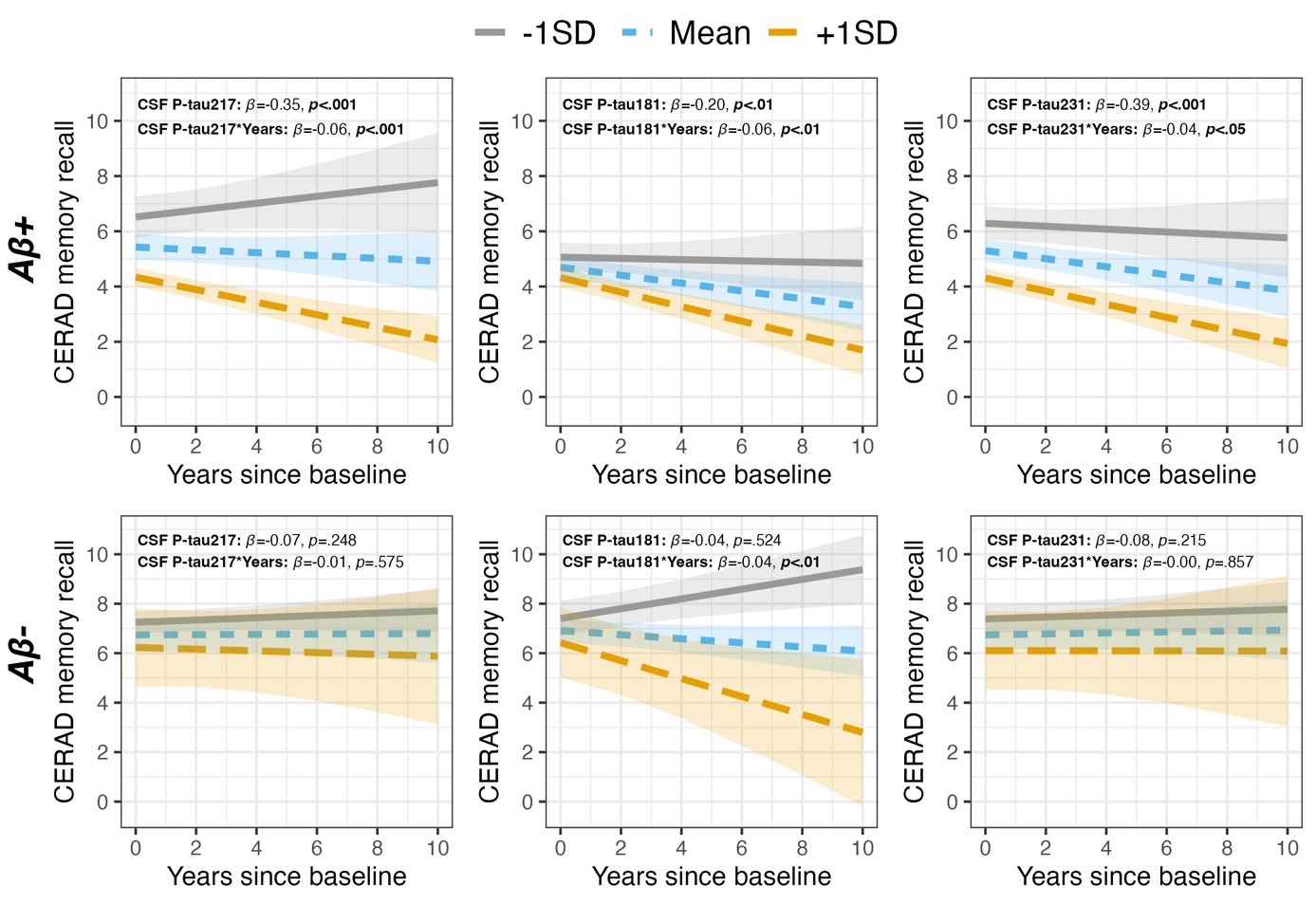

**Supplementary figure 4 (S4).** **Baseline and longitudinal associations of CSF p-tau markers with the Consortium to Establish a Registry for Alzheimer’s Disease (CERAD) memory recall test in cohort-1**. **Figure S4A-C)** show the baseline and longitudinal associations of CSF p-tau217, p-tau181 and p-tau231 with the CERAD memory recall test in Aβ+ individuals. **Figure S4D-F)** show the baseline and longitudinal associations of CSF p-tau217, p-tau181 and p-tau231 with the CERAD memory recall test in Aβ- individuals. The lines display associations between the biomarker at −1SD (grey), Mean (blue) and +1SD (orange) and the dependent variable at baseline and over time.

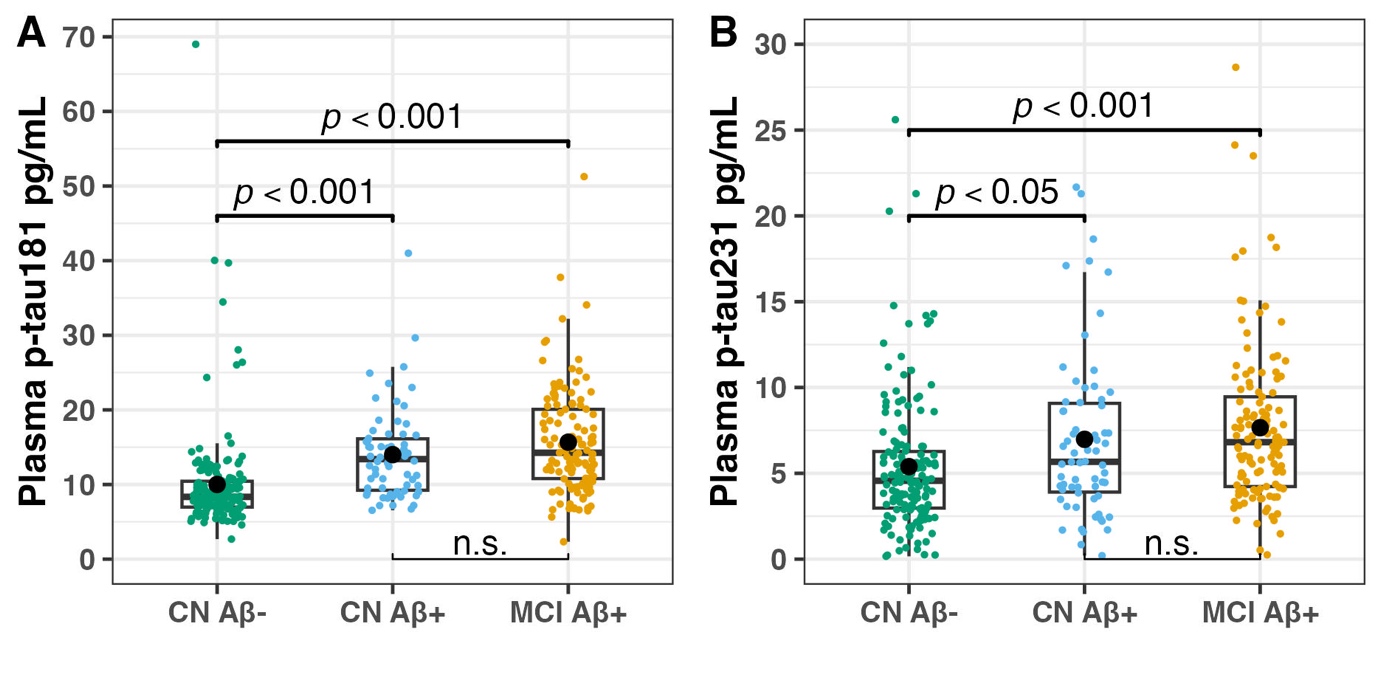

**Supplementary figure 5 (S5).** **Plasma P-tau181 and P-tau231** **concentrations by cognitive status in cohort 1.** Boxplots showing concentrations of plasma p-tau181 and p-tau231 (pg/ml) in Cognitively normal (CN) Aβ-, CN Aβ+ and Mild Cognitive Impairment (MCI) Aβ+ individuals in Dementia Disease Initiation (cohort1). The brackets show statistically significant differences between the groups (FDR adjusted p-values).
